# Earlier confirmation of glaucoma progression with risk-weighted visual field and OCT scheduling

**DOI:** 10.64898/2026.11.16.26368187

**Authors:** Liyin Chen, Asahi Fujita, Mousa Moradi, Mohammad Eslami, Tobias Elze, Nazlee Zebardast

## Abstract

Glaucoma is monitored with visual field (VF) tests of function and optical coherence tomography (OCT) of structure, but models that predict progression do not determine when each test should be repeated. Here we present GEAR (gated expected-delay allocation rule), a policy that pairs a longitudinal risk predictor with a decision layer and gives each eye separate VF and OCT intervals. In the decision layer, clinical gates define the admissible test intervals, and an explicit objective selects the intervals that minimize risk-weighted detection delay plus a penalty on expected testing, with penalties fixed to a routine-testing budget. In retrospective replay of held-out records (5,935 visits from 1,681 patients), GEAR with risk-based confirmation timing confirmed simulated progression 3.4 months earlier than annual VF and OCT testing (95% confidence interval, 2.7-4.1) at a comparable testing rate. The reduction persisted against a guideline-informed comparator and a reconstructed Kalman comparator, under an alternative onset assumption and with a VF-only adaptation in an external dataset.

## Introduction

Monitoring a chronic eye disease requires a decision about when to repeat each test. Progression risk estimates describe the probability of a defined change over time, but a monitoring decision also depends on the consequences of waiting and the cost of testing. When function and structure are measured by different tests, their intervals need not coincide. A decision layer placed on top of a risk predictor can make this translation explicit by defining the allowed schedules, scoring their delay-testing trade-offs and selecting among them under a stated budget.

Glaucoma poses this allocation problem in a clear form. Visual field (VF) testing measures function, while optical coherence tomography (OCT) assesses structure. Repeated measurements are needed to distinguish progression from variability, and their timing affects when change can be recognized [1,2]. Increasing testing frequency can shorten detection time but also increases the number of examinations. The decision is therefore not only how often to monitor an eye, but how to distribute functional and structural testing according to their separate risks and clinical usefulness.

Personalized monitoring has a substantial methodological history. Variable VF intervals with prompt confirmation of suspected deterioration and clustered testing strategies have been investigated in simulation [3,4]. Schell and Helm used filtered forecasts of VF indices and intraocular pressure to time the next examination for each patient [5,6], while Kazemian and colleagues extended this approach to select which tests to take in each six-month period (none, intraocular pressure alone or intraocular pressure with a VF) together with a target intraocular pressure [7]. Recent work has also examined staged VF testing, with initially intensive measurement followed by reduced frequency [8]. Outside ophthalmology, a policy learned by reinforcement learning has set personalized mammography screening intervals from an image-based risk model [9]. These studies establish that a forecast of progression can set a monitoring interval, and that such intervals can identify progression sooner than fixed schedules with a similar number of examinations [5,6].

This body of work leaves four gaps. First, the forecasting-based schedules were developed on VF indices and intraocular pressure from trial cohorts examined at six-month visits, and none included structural imaging [5–7], while the screening policy schedules a single test [9]. OCT is now part of routine glaucoma monitoring, and VF and OCT differ in when they can show change [1,2], yet to our knowledge no prediction-informed schedule assigns the two tests separate intervals. Second, the balance between detection delay and testing burden has been set indirectly, through progression thresholds and confidence levels [5,6], through cost weights tied to a clinician-selected level of aggressiveness [7] or through a policy learned from a reward that combines early detection with screening cost [9]. These settings do not by themselves state which schedules are clinically admissible for an eye, when no recommendation should be made or what average testing budget the schedule is meant to meet. Third, multimodal models can forecast future glaucoma worsening from early VF, OCT and clinical data [10], and longitudinal imaging models can estimate time-varying ophthalmic risk [11], but these forecasts stop at a risk estimate, and prognostic discrimination alone does not determine when each test should be repeated. Fourth, these schedules have been evaluated by simulation or retrospective analysis, and to our knowledge none has been tested for its effect on the intervals that clinicians choose. The open question is how to translate separate risk curves for a functional and a structural test into separate intervals while retaining clinically admissible choices, an explicit objective and a specified testing budget, and whether the resulting schedule helps the clinician who uses it.

Here we present GEAR (gated expected-delay allocation rule), a policy that schedules VF and OCT testing for eyes monitored for glaucoma. At a clinic visit, GEAR outputs separate windows for the next VF and OCT tests for an eye, optimized toward catching disease progression sooner while taking testing burden into account. GEAR has two parts. A longitudinal risk predictor estimates, from the eye’s VF, OCT and clinical history, the probability that progression becomes detectable on each test over the next two years. A decision layer then converts these risk curves into test intervals, and it has a clinically grounded component and a mathematically optimized component. The clinically grounded component is a set of clinical gates that determine whether a recommendation can be made and which pairs of VF and OCT intervals are admissible. The mathematically optimized component selects among the admissible pairs by minimizing risk-weighted detection delay plus a penalty on expected testing, with penalties fixed on development data to meet an average routine-testing budget. When a test shows a possible change, the decision layer also sets how soon that test is repeated. We evaluated GEAR in retrospective replay of held-out glaucoma records against annual testing, a guideline-informed comparator and a reconstructed Kalman comparator. We then examined its sensitivity to assumptions about progression onset and confirmation, a later-period refit and a VF-only adaptation in an external dataset.

## Results

### GEAR pairs a longitudinal risk predictor with a clinically gated decision layer

GEAR turns the test history of an eye into a monitoring schedule in two stages (Fig. 1). A risk predictor first estimates, for each test used to monitor the disease, the cumulative probability that progression becomes detectable on that test over a set of future horizons (Fig. 1a). A decision layer then converts these risk curves into an interval for each test, and it has two components.

**Fig. 1.**
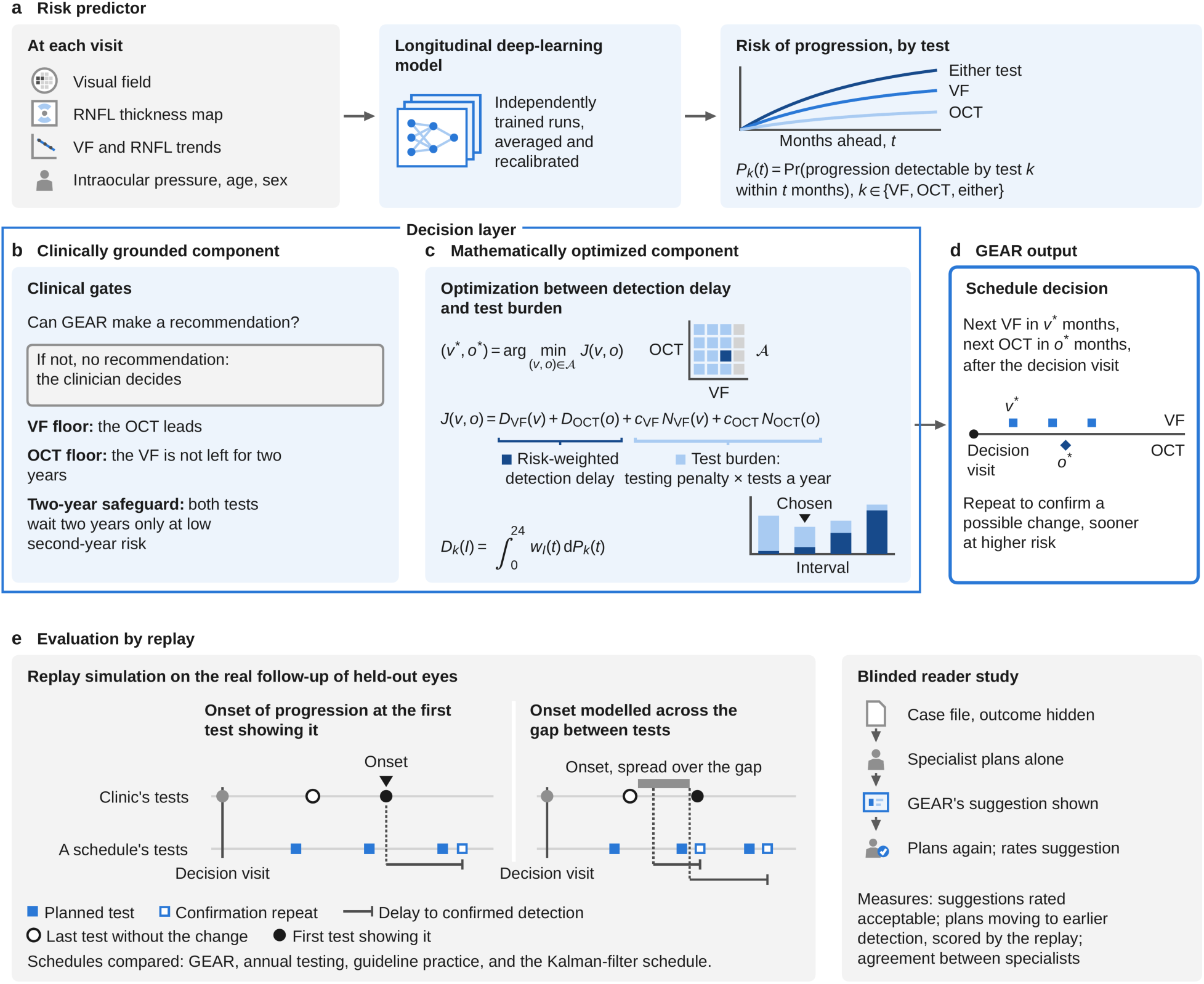
GEAR pairs a longitudinal risk predictor with a clinically gated decision layer. a, Risk predictor. A longitudinal deep-learning model reads up to eight test days of VF tests, OCT RNFL thickness maps and summaries, intraocular pressure, age and sex. Ten runs are averaged and recalibrated to give the cumulative risk of detectable progression on VF, on OCT and on either test. The either-test risk also counts major glaucoma operations. b, Clinically grounded component of the decision layer. Eligibility gates return no recommendation when recent reliable tests are missing, and a second route does so when both tests are at their measurement floor or the ten runs split evenly on the two-year safeguard. Measurement-floor gates apply at an MD of −20 dB or worse, an RNFL thickness of 57 µm or less or an OCT that cannot assess progression. The two-year safeguard allows both tests at 24 months only if either-test risk rises by at most 1.5 percentage points in the second year and the OCT is above its floor. The remaining interval pairs are admissible. c, Mathematically optimized component. The delay-testing objective J adds, for each test, the risk-weighted detection delay and a testing penalty multiplied by the expected tests per year, and the admissible pair with the smallest J is selected (Methods, equations (1) and (2)). Testing penalties were fixed on the development set so that routine testing averaged two tests per eye per year, with repeats additional. d, GEAR output. A test showing a possible change is repeated after 1 month when six-month either-test risk is at least 1.4% and after 2 months otherwise. e, Evaluation by replay. Each schedule is laid over the same recorded follow-up of test-set eyes. Observed-onset replay places onset at the first test showing progression, and interval-onset replay spreads onset evenly across the gap since the last test without the change. Comparators repeat 2 months after a possible change, and operations do not count as confirmed progression. Risk curves, bars and timelines are illustrative. GEAR, gated expected-delay allocation rule; VF, visual field; OCT, optical coherence tomography; RNFL, retinal nerve fiber layer; MD, mean deviation.

The clinically grounded component applies clinical gates, which determine whether a recommendation can be issued and which combinations of intervals are admissible (Fig. 1b). The mathematically optimized component scores every admissible combination by its risk-weighted detection delay plus a penalty on expected testing, and selects the combination with the lowest score (Fig. 1c). The decision layer accepts risk curves from any predictor, provided that the probabilities refer to the outcomes and horizons it uses and are calibrated.

For glaucoma, the two tests were VF and OCT. The risk predictor was a longitudinal deep-learning model that used VF measurements, OCT retinal nerve fiber layer (RNFL) thickness maps and summaries, intraocular pressure, age and sex (Fig. 1a). It was trained on three progression labels. The VF label required agreement of at least two eligible progression algorithms, and the OCT label required confirmed RNFL thinning. The either-test label combined the two and additionally counted major glaucoma operations as events. At each visit, GEAR outputs separate windows for the next VF and OCT tests for an eye, optimized toward catching disease progression sooner while taking testing burden into account (Fig. 1d). Each window is one of four intervals (4, 6, 12 or 24 months) after which the next test of that kind is due.

We developed and evaluated GEAR in records from Mass Eye and Ear (2016-2025), with one patient in five assigned to the test set and the remainder to the development set. The unit of analysis was the decision visit, defined as a test day (a day with a VF or an RNFL thickness map) at which GEAR was asked for a schedule. Each eye contributed up to five decision visits (its study entry and its next four test days), and each visit used only the tests recorded on or before that day. The development set comprised 47,834 decision visits from 16,873 eyes of 10,662 patients, and the test set comprised 12,260 decision visits from 4,250 eyes of 2,696 patients (Table 1). Unless stated otherwise, results refer to the test set.

**Table 1.** Characteristics of the development and test sets.

| **Characteristic** | **Development set**<br>10,662 patients, 16,873 eyes, 47,834 decision visits | **Test set**<br>2,696 patients, 4,250 eyes, 12,260 decision visits |
|---|---|---|
| Age (years), median (interquartile range) | 65 (55 to 73) | 65 (54 to 73) |
| Women | 58% | 58% |
| MD (dB), median (interquartile range) | -1.5 (-3.6 to -0.2) | -1.6 (-3.6 to -0.2) |
| MD stage, mild / moderate / severe | 86% / 8.7% / 4.9% | 86% / 8.6% / 5.0% |
| RNFL (µm), median (interquartile range) | 83 (73 to 91) | 83 (73 to 91) |
| Follow-up (months), median (interquartile range) | 31 (13 to 54) | 31 (13 to 54) |

Values describe decision visits. Age is taken at the visit, and women are counted once per patient. MD is the mean deviation of the latest reliable VF on or before the visit and is staged as mild (above -6 dB), moderate (-12 to -6 dB) or severe (below -12 dB). RNFL is the latest average retinal nerve fiber layer thickness. Follow-up runs from the visit to the eye's last reliable VF or recorded OCT scan before any glaucoma operation. One eye contributes up to five decision visits. The study holds the decision visits whose latest OCT on or before the visit has an RNFL thickness map.

The decision layer received recalibrated risk at five horizons from 3 to 24 months and considered the 16 pairs of 4-, 6-, 12- or 24-month VF and OCT intervals. Its clinically grounded component applied three kinds of clinical gate (Fig. 1b). Eligibility gates withheld a recommendation when the latest VF was unreliable, when fewer than two reliable VFs existed or when the latest reliable VF or OCT was more than two years old or absent. Measurement-floor gates applied when one test could no longer show further change, and they kept the other test at 12 months or sooner. A two-year safeguard allowed both tests to be deferred for 24 months only when predicted either-test risk rose by at most approximately 1.5 percentage points during the second year. The remaining conditions for no recommendation are given in the Methods.

Among admissible pairs, the mathematically optimized component selected the pair that minimized the sum of modality-specific risk-weighted detection delays and penalties for expected test counts (Fig. 1c and Methods, equations (1) and (2)). The testing penalties were selected on development patients to meet an average budget of two routine tests per eye per year, with confirmation repeats additional, and then fixed. This budget governed population-average routine testing during development selection rather than imposing the same interval or a hard test-count limit on every eye.

A test can suggest progression without yet meeting the confirmation criterion. After such a possible change, GEAR scheduled a repeat of the same test to confirm it (Fig. 1d). The repeat came after 1 month when predicted six-month either-test risk was at least approximately 1.4% and after 2 months otherwise, whereas every comparator repeated after 2 months. The delays and test counts reported below therefore reflect routine intervals and confirmation timing together.

The risk predictor discriminated the two-year composite of progression or major glaucoma operation with an area under the receiver operating characteristic curve of 0.83 (95% confidence interval (CI), 0.80-0.86) at 7,672 visits (Supplementary Fig. 1). We evaluated GEAR by replay, in which each schedule was laid over the recorded follow-up of the same eyes and the time from progression onset to its confirmation was simulated (Fig. 1e). We compared GEAR with annual VF and OCT testing, a guideline-informed comparator based on disease stage and intraocular pressure, and a reconstruction of a published Kalman-filter VF schedule [5,6] paired with annual OCT (the reconstructed Kalman comparator). Replay counted progression confirmed on VF or OCT and did not count operations, because no schedule can detect an operation sooner. It included 5,935 scored visits from 2,601 eyes of 1,681 patients with sufficient outcome follow-up, and confirmation delay was evaluated at the 371 scored visits that were followed by progression within two years (progression-associated visits; Supplementary Table 1).

### GEAR confirms simulated progression earlier at comparable aggregate testing

GEAR reduced mean simulated confirmation delay from 10.0 months with annual VF and OCT testing to 6.6 months, a paired reduction of 3.4 months (95% CI, 2.7-4.1; Fig. 2a,b and Supplementary Table 2). Total testing, including confirmation repeats, was comparable at 2.17 versus 2.18 tests per eye per year. GEAR scheduled 1.97 routine tests per eye per year, and with repeats its total comprised 1.28 VFs and 0.89 OCT scans.

**Fig. 2.**
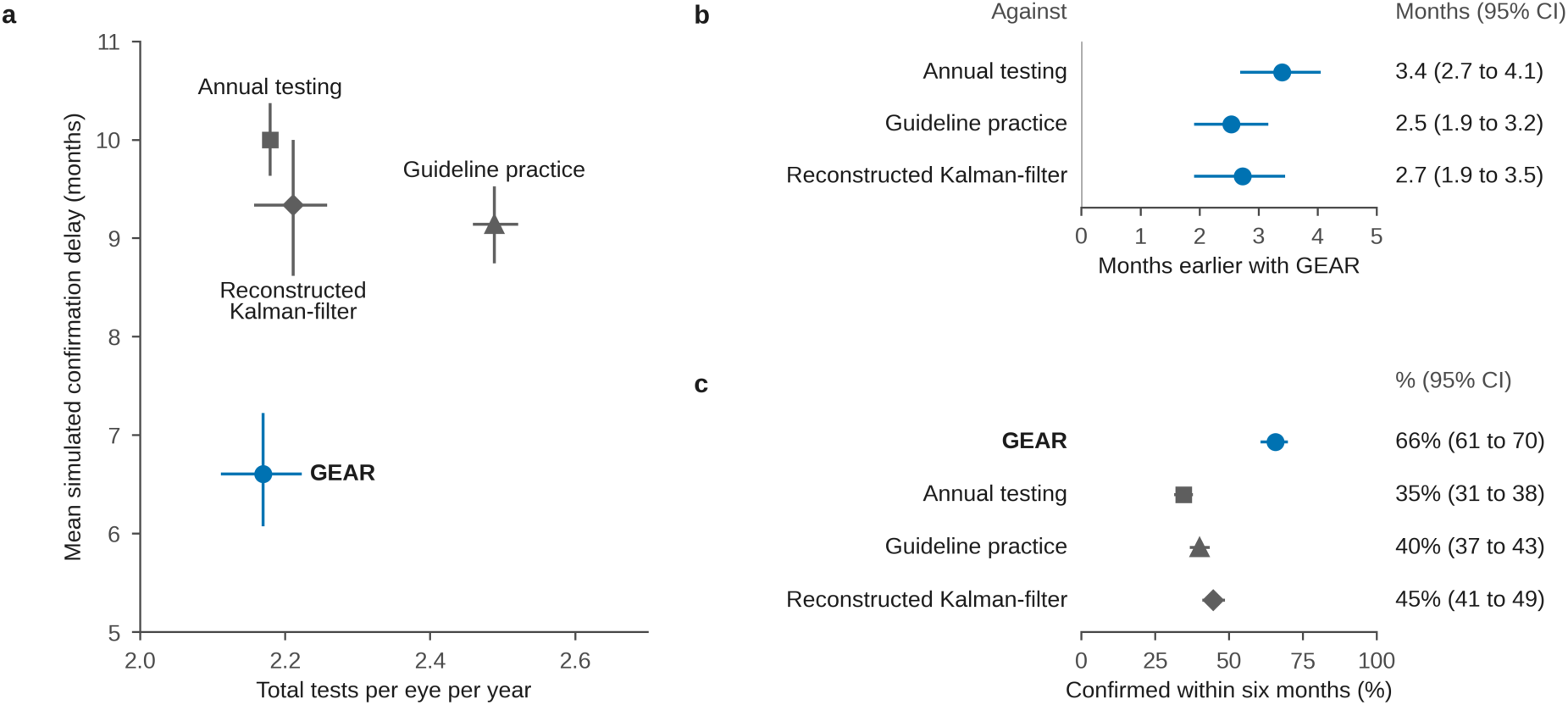
Earlier simulated confirmation with GEAR at comparable aggregate testing. a, Mean simulated confirmation delay versus total expected tests per eye per year for GEAR (blue circle) and for annual testing (gray square), the guideline-informed comparator (gray triangle) and the reconstructed Kalman comparator (gray diamond). Delay is evaluated at 371 progression-associated decision visits from 201 eyes of 183 patients; testing frequency is averaged over all 5,935 scored visits from 2,601 eyes of 1,681 patients. Horizontal and vertical whiskers show marginal 95% confidence intervals (CIs). b, Paired delay reductions, calculated as comparator delay minus GEAR delay; positive values indicate earlier confirmation with GEAR. c, Percentage of progression-associated visits confirmed within six months, with 95% CIs. All panels use observed-onset replay. Testing counts include VF and OCT examinations and confirmation repeats. The reconstructed Kalman comparator forecasts MD, PSD and intraocular pressure, with a confidence-region size of 0.5, and is paired with annual OCT. CIs are from 1,000 patient-level bootstrap resamples with the risk predictor and decision-layer settings held fixed. VF, visual field; OCT, optical coherence tomography; MD, mean deviation; PSD, pattern standard deviation.

Mean confirmation delay was 9.1 months with the guideline-informed comparator and 9.3 months with the reconstructed Kalman comparator. Corresponding reductions with GEAR were 2.5 months (95% CI, 1.9-3.2) and 2.7 months (1.9-3.5), at comparator testing rates of 2.49 and 2.21 tests per eye per year (Supplementary Fig. 2). GEAR thus achieved earlier simulated confirmation at approximately the same aggregate testing frequency as annual testing and the reconstructed Kalman comparator, and with 13% fewer tests than the guideline-informed comparator.

The first positive examination dates the observation of change, although detectable progression may have begun between examinations. Observed-onset replay places onset at that first positive test. To assess sensitivity to this timing assumption, interval-onset replay distributed onset uniformly between the last negative and first positive examinations. Under this alternative, GEAR shortened simulated confirmation by 3.6 months (95% CI, 2.8-4.3) relative to annual testing, 2.6 months (2.0-3.3) relative to the guideline-informed comparator and 3.1 months (2.2-3.9) relative to the reconstructed Kalman comparator (Supplementary Fig. 3 and Supplementary Table 2).

In observed-onset replay, progression was confirmed within six months at 66% of progression-associated visits (95% CI, 61-70%), compared with 35% (31-38%) under annual testing, 40% (37-43%) with the guideline-informed comparator and 45% (41-49%) with the reconstructed Kalman comparator (Fig. 2c). The paired increase over annual testing was 31 percentage points (95% CI, 26-36).

### Testing differs by modality and follows predicted risk

We next examined how GEAR distributed testing between modalities and across levels of predicted risk (Fig. 3). Across all 12,260 decision visits, a 24-month interval was assigned more often for OCT (44%) than for VF (29%). Both tests remained annual at 7.8% of visits, both were deferred for 24 months at 19.7%, and at least one was scheduled at 4 or 6 months at 24% (Fig. 3a and Supplementary Tables 3 and 4).

**Fig. 3.**
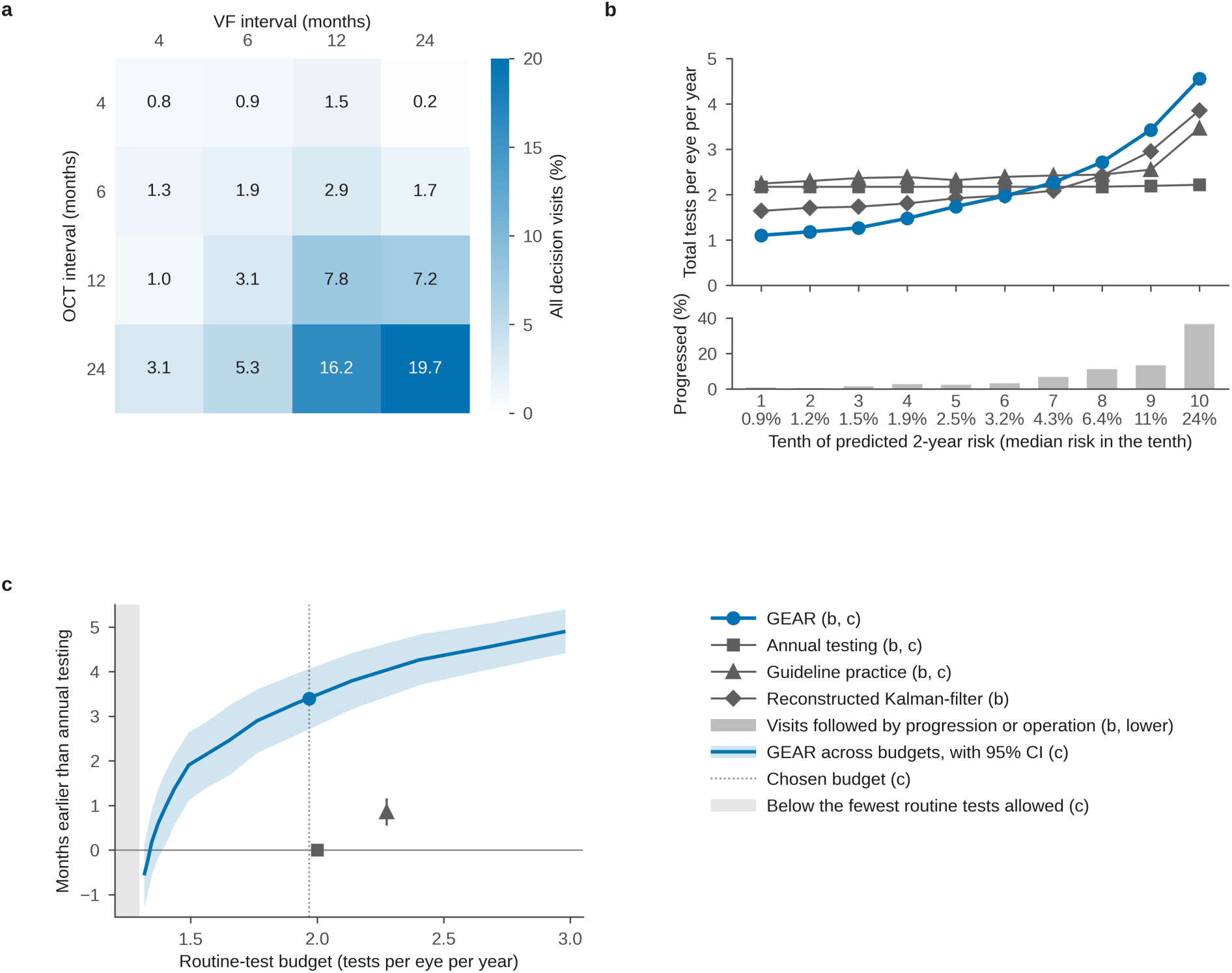
Allocation of VF and OCT testing by GEAR across modalities and predicted risk, and its dependence on the testing budget. a, Joint VF/OCT interval allocation among 12,260 decision visits from 4,250 eyes of 2,696 patients. VF intervals run across columns and OCT intervals down rows. Cell percentages use all decision visits as the denominator; the 16 cells account for 9,154 visits (74.7%), and 3,106 visits (25.3%) received no recommendation. The white-to-blue scale indicates the percentage of all visits. The grid shows selected schedules, not a universal admissibility mask. b, Total expected tests per eye per year scheduled in each tenth of the predicted two-year risk of progression or a major glaucoma operation over the 5,935 scored visits (top; markers follow Fig. 2), and the percentage of visits in each tenth followed within two years by progression or a major glaucoma operation, among the visits at which progression could be judged (bottom). The label under each tenth gives its median predicted risk. c, Months earlier with GEAR than with annual testing in the observed-onset replay as both testing penalties of GEAR are scaled together, plotted against the routine-test budget that each scale produces (mean routine tests per eye per year over all scored visits, repeats excluded), with the 95% confidence interval (CI) shaded. The dotted line marks the budget selected on development patients (2.00 routine tests per eye per year there; 1.97 at these visits). Annual testing and the guideline-informed comparator are shown at their own routine-test rates, the comparator with its 95% CI. The shaded region lies below the fewest routine tests any admissible schedule allows (1.30). Intervals are from 1,000 patient-level bootstrap resamples. VF, visual field; OCT, optical coherence tomography.

GEAR shifted testing from visits at lower predicted risk to visits at higher predicted risk (Fig. 3b). Across tenths of the predicted two-year risk of progression or a major glaucoma operation, the testing scheduled by GEAR rose from 1.10 tests per eye per year in the lowest tenth to 4.55 in the highest. Annual testing scheduled 2.17 to 2.22 tests per eye per year in every tenth, and the guideline-informed comparator 2.24 to 2.55, rising to 3.46 in the highest tenth. The reconstructed Kalman comparator, which times the VF from forecasts of MD, also scheduled more tests at higher predicted risk, from 1.64 to 3.85 tests per eye per year. The share of visits followed within two years by progression or a major glaucoma operation rose from 1.0% in the lowest tenth to 37% in the highest. Testing was therefore more frequent at the 443 visits followed by observed progression or an operation, at which GEAR scheduled 3.65 tests per eye per year, compared with 2.19 for annual testing, 2.98 for the guideline-informed comparator and 3.01 for the reconstructed Kalman comparator.

The routine-test budget set the size of GEAR’s advantage (Fig. 3c). When both testing penalties were scaled together, the advantage over annual testing grew with the routine-test budget, and its 95% CI excluded zero from about 1.4 routine tests per eye per year upward. At the selected budget it was 3.4 months. No admissible schedule used fewer than 1.30 routine tests per eye per year. Even with perfect foresight of progression, spending GEAR’s total with every progressing visit at its largest admissible testing rate would have left 1.86 tests per eye per year at visits without observed progression.

### GEAR’s advantage in earlier detection persists across subgroups, alternative assumptions and a later period

To check whether the delay reduction was confined to particular eyes or depended on the assumptions of the replay, we performed extensive sensitivity analyses. Estimated confirmation delays were shorter with GEAR than with annual testing and the guideline-informed comparator across the evaluated disease stages, age groups, sexes and calendar periods (Fig. 4a and Supplementary Table 5). Reductions relative to annual testing were 2.1, 4.9 and 7.0 months in mild, moderate and severe disease, respectively, and corresponding reductions relative to the guideline-informed comparator were 2.0, 4.7 and 1.9 months. Among eyes losing at least 1 dB of VF mean deviation (MD) per year, the comparison was based on 36 progression-associated visits and favored GEAR by 3.2 months over annual testing (95% CI, 1.1-4.7). Progression-speed groups were defined from follow-up and were available for 258 of the 371 visits. Interval-onset analyses showed similar patterns (Supplementary Fig. 4). These estimates describe within-group comparisons; differences between groups were not tested.

**Fig. 4.**
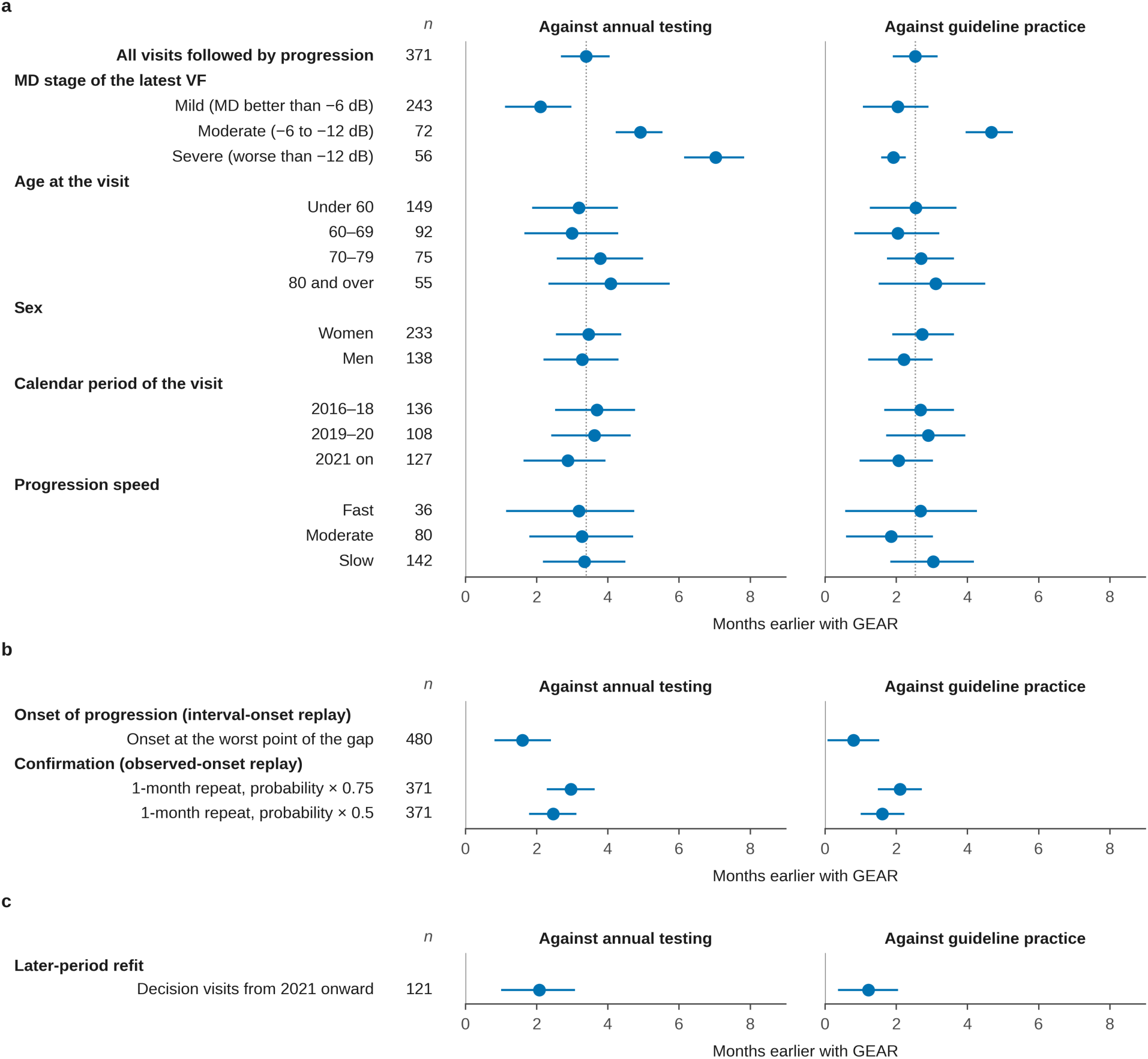
Subgroup estimates and sensitivity analyses including later-period evaluation. a, Paired confirmation-delay reductions for GEAR relative to annual testing (left) and the guideline-informed comparator (right) in observed-onset replay; n denotes progression-associated decision visits. The overall 371-visit row is a reference. In a, the dotted lines mark the overall estimates. MD stage is based on the latest VF at the decision visit and is mild (MD above-6 dB), moderate (−12 to −6 dB) or severe (MD below −12 dB). Age and calendar period are defined at the decision visit. Calendar subgroups use the main risk predictor fitted across all years. Progression speed is defined from the follow-up MD trend, requiring at least five VFs over at least two years, and is fast (loss of at least 1 dB per year), moderate (0.3 to less than 1 dB per year) or slow (loss below 0.3 dB per year or an improving trend). A slope was available for 258 of 371 visits. b, Sensitivity to adverse onset placement and reduced one-month confirmation success. Adverse placement uses the interval-onset analysis points at 480 visits in 230 eyes and selects each eye’s onset to minimize the gain of GEAR over the comparator. One-month confirmation success is multiplied by 0.75 or 0.50, with the two-month probability unchanged, in the 371-visit observed-onset population. c, Later-period evaluation after refitting the risk predictor on development visits through 31 December 2018 with outcomes confirmed by 31 December 2020, together with the no-recommendation rule, repeat-risk threshold and two-year safeguard; other components were retained as specified in Methods. Positive differences indicate earlier simulated confirmation with GEAR. Whiskers show paired 95% CIs from 1,000 patient-level bootstrap resamples; adverse onset placements are held fixed within resamples. No between-group or interaction tests were performed. MD, mean deviation; VF, visual field.

The delay advantage persisted when onset was placed, for each eye, at the point within its inter-test interval that minimized the gain of GEAR over each comparator. This analysis used the 480-visit interval-onset population and yielded reductions of 1.6 months versus annual testing (95% CI, 0.8-2.4) and 0.8 months versus the guideline-informed comparator (0.1-1.5; Fig. 4b). Allowing onset to be up to eight times more likely in some months of the interval than others also retained positive delay reductions (Supplementary Table 6).

To estimate confirmation success at one month, we reduced that success probability to 75% or 50% of the probability at two months. In the 371-visit observed-onset population, estimated gains over annual testing were then 3.0 and 2.5 months, respectively, and those over the guideline-informed comparator were 2.1 and 1.6 months (Fig. 4b). These analyses test sensitivity to early-repeat effectiveness.

Two controls examined whether the reduction came from eye-specific risk or from the number of tests alone. Shuffling predicted risks between visits increased confirmation delay by 4.7 months relative to GEAR (95% CI, 3.8-5.4), with a testing frequency of 2.33 tests per eye per year. A uniform schedule of both tests every 12.1 months matched the total frequency of GEAR (2.17 tests per eye per year) but confirmed progression 3.5 months later (95% CI, 2.7-4.1).

The budget of GEAR could be moved without changing the risk predictor or the gates. Re-selecting the two testing penalties on the development set for routine budgets of 1.5, 2.5 and 3.0 tests per eye per year gave mean simulated confirmation delays of 8.4, 5.6 and 5.1 months, compared with 6.6 months at 2.0 (Supplementary Table 7).

We also tested whether a more complex policy that learns the mapping from risk to intervals would outperform the explicit objective. A learned-policy comparator received the same inputs, clinical gates, repeat rule and development-selected budget as GEAR and differed only in how it chose among the admissible interval pairs. It did not measurably shorten simulated confirmation relative to GEAR (6.50 versus 6.60 months; difference 0.10 months; 95% CI, −0.33 to 0.54), while using 0.07 more tests per eye per year (0.04-0.10; Supplementary Table 8). Its routine testing rose from 1.98 tests per eye per year on the development set to 2.03 on the test set, whereas that of GEAR was 1.98 and 1.97. The two policies chose the same interval pair at 45% of visits with a recommendation, so different schedules reached a similar delay-testing trade-off. When one fold of the development patients was withheld and each policy was refitted (a post hoc check), the comparator kept its chosen pair at 69% of visits on average and GEAR at 91%.

To assess later-period performance at the same hospital, we refitted the risk predictor and the decision-layer thresholds using development visits through 2018 and outcomes confirmed by 2020, then evaluated visits from 2021 onward (Fig. 4c; Methods). Delay estimates used 121 progression-associated visits from 76 eyes, and testing rates used 1,962 scored visits. GEAR confirmed progression 2.1 months earlier than annual testing (95% CI, 1.0-3.1), 1.2 months earlier than the guideline-informed comparator (0.4-2.1) and 1.8 months earlier than the similarly refitted Kalman comparator (0.6-2.9). Interval-onset reductions were 2.3, 1.3 and 2.1 months, respectively.

### A VF-only adaptation of GEAR shortens simulated confirmation in an external dataset

We evaluated a VF-only adaptation, GEAR-VF, in the University of Washington’s public perimetry dataset [12]. Because OCT and intraocular pressure data were unavailable, GEAR-VF paired the VF terms of the decision layer, the VF measurement floor and risk-based confirmation timing with a separately developed Cox model of VF summaries and age that was trained and recalibrated on our development patients. The evaluation included 6,235 scored visits from 1,875 eyes of 1,025 patients, out of 18,342 decision visits. Observed-onset delay was evaluated at 435 progression-associated visits in 231 eyes of 194 patients. Simulated confirmation occurred 2.8 months earlier than with annual testing (95% CI, 2.4-3.1), 1.7 months earlier than with the stage-based guideline-informed comparator (1.4-2.0) and 1.2 months earlier than with the reconstructed Kalman comparator (0.8-1.7), which here forecast MD and pattern standard deviation (PSD). Interval-onset reductions were 2.8, 1.7 and 1.5 months, respectively (Supplementary Fig. 5 and Supplementary Table 9). These findings support transfer of the decision layer to a separately developed Cox model and independent perimetry data, with earlier simulated confirmation than all three comparators.

## Discussion

We developed GEAR, a policy that converts test-specific progression risk into separate VF and OCT intervals for glaucoma monitoring under a development-selected testing budget. A risk predictor supplies the risk curves, and a decision layer converts them into intervals. Within the decision layer, clinical gates define the admissible schedules and an explicit delay-testing objective selects among them. In retrospective replay, GEAR shortened simulated confirmation by 3.4 months relative to annual VF and OCT testing at comparable aggregate test counts. The advantage persisted under an alternative onset assumption, population stratification sensitivity analysis and external validation.

Separating the decision layer from the risk predictor makes the operation of GEAR inspectable. A change in a risk curve changes the modeled cost of waiting, a clinical gate removes an inadmissible schedule and the testing penalties express the selected resource trade-off. Re-selecting these two penalties moves GEAR to a different testing budget without refitting the risk predictor or altering the gates. Each recommendation also carries one reason per test, which names the gate or the side of the objective that set the interval (Methods). The observed allocation patterns show how these choices distribute testing across modalities and levels of predicted risk.

The delay reduction was not uniform across disease stages. Relative to annual testing it was 2.1 months in mild disease and 4.9 and 7.0 months in moderate and severe disease, although differences between stages were not tested and the reduction relative to the guideline-informed comparator in severe disease was 1.9 months. Earlier confirmation may matter most in the moderate and severe eyes in which GEAR shortened it most.

The later-period and external analyses showcase different aspects of applicability. The internal later-period refit retained an advantage over annual testing and used earlier data to refit the risk predictor and the decision-layer thresholds rather than every component. The external evaluation used a separately developed VF-only Cox model and an independent perimetry dataset, and it supports use of the decision layer with a different predictor and input set. GEAR-VF shortened simulated confirmation relative to all three comparators. These findings support adaptation of the decision layer; external multimodal performance remains untested. The formulation is not tied to glaucoma. Other chronic eye diseases are also followed with a functional and a structural test, as in annual screening for hydroxychloroquine retinopathy with automated VF and OCT [13]. Monitoring of this kind generally involves the elements that the decision layer encodes, namely a separate risk of detectable change on each test, confirmation of a suspected change before acting and limits on what each test can measure.

In glaucoma, a decision layer placed on top of a risk predictor translated separate VF and OCT risk forecasts into explicit monitoring schedules whose delay-testing trade-off could be evaluated. The formulation specifies the risk for each test, defines clinically admissible schedules and chooses intervals using an explicit objective with a development-selected budget, and it could be specified for other chronic eye diseases that are monitored with paired functional and structural tests. The reported evidence supports GEAR in retrospective glaucoma replay and motivates prospective evaluation of its use in clinical monitoring.

## Methods

### Data and study design

This retrospective study used the records of Mass Eye and Ear, Boston, Massachusetts, USA. They held Humphrey Field Analyzer VFs (24-2 tests, and 30-2 tests reduced to their 24-2 points; SITA Standard, SITA Fast and SITA Faster), Cirrus OCT RNFL thickness (average, eight sectors and the thickness map), intraocular pressure, age and sex, from clinic visits from 2016 to March 2025. A VF was reliable when its false-positive rate was 0.20 or less, its false-negative rate 0.20 or less (recorded, for SITA Standard and Fast tests) and its fixation losses 0.33 or less where recorded, with an MD between −40 and +5 dB. An eye entered the study at the first test day on or after the latest of its second reliable VF, its first thickness map and its first clinic record. Eyes with a glaucoma operation on or before entry were left out. Patients had been assigned to five folds by a stratified patient-level split made before this project; one fold (one patient in five) formed the test set and the other four formed the development set, with all of a patient’s eyes in one fold. Every setting of the risk predictor and of the decision layer fitted in this study was fitted on development patients.

### Ethics

The Institutional Review Board at Mass General Brigham approved this retrospective study, conducted in accordance with the Declaration of Helsinki. Given the retrospective design, the requirement for informed consent was waived.

### Decision visits and the study population

The decision visits of an eye were its entry and its next four test days (days with a VF, reliable or not, or a thickness map), one per calendar day. At each, the latest test of each kind on or before the visit was read. The study held the decision visits whose latest OCT on or before the visit had an RNFL thickness map. Of the 9,154 test-set decision visits with a recommendation, the 5,935 followed for 2 years or until progression or a major glaucoma operation were scored.

### Progression outcome

Replay progression was defined as worsening confirmed on the VF or OCT, whichever was confirmed first. Separate VF and OCT labels were used for modality-specific risk prediction. VF worsening required at least 2 of the 6 methods compared by Saeedi et al. [14] to agree, among those able to judge the eye. The trend methods were the MD and visual field index slopes (−1 dB or −1% per year or worse, one-sided P < 0.05), pointwise linear regression (at least 3 of 52 points with slopes of −1 dB per year or worse, P < 0.01) and permutation analysis of pointwise linear regression (PoPLR; P < 0.05 over 5,000 permutations) [15], each needing at least 5 VFs over at least 2 years, 2 of them after entry. The event methods were the AGIS and CIGTS scores, a rise of at least 4 and 3 points on 3 consecutive later VFs, not judged above a baseline score of 16 and 17. In the outcome only, the MD and visual field index of SITA Faster tests were shifted by the measured difference from SITA Standard in three damage bands. The baseline was the latest reliable VF on or before entry; worsening was confirmed on the day two methods agreed, and its onset was the earliest onset among them. An eye whose starting VF had an MD of −20 dB or worse was not judged on the VF. OCT worsening followed the rule of the COAST trial [16], which requires average RNFL thinning of at least 10 µm from the baseline scan (the latest scan on or before entry) on two consecutive scans, the first marking the onset and the second the confirmation. Eyes with a baseline of 67 µm or less were not judged on the OCT. A scan counted when its signal strength was 6 or more and its average thickness between 30 and 150 µm, keeping the highest signal on a day. The onset of either-test progression was the earlier of the two onsets. The risk predictor’s either-test labels also counted a major glaucoma operation (trabeculectomy, tube or cyclophotocoagulation) as an event; the replay never did, because no schedule can find an operation sooner.

### Risk predictor

The risk predictor was a longitudinal deep-learning model that gave the risk that progression would become detectable on the VF, on the OCT and on either test within 3, 4, 6, 12, 24 and 36 months (Supplementary Fig. 1). The decision layer used the five horizons up to 24 months. For each decision visit it read up to 8 calendar days with a test on or before the visit, newest first. Each day held a summary of the VF (256 numbers) from a VF encoder and a summary of the RNFL thickness map (512 numbers) from a map encoder (RNFLT2Vec), both trained earlier without progression labels and kept fixed; 9 context values (MD, PSD and visual field index, average RNFL thickness, the VF’s fixation-loss, false-positive and false-negative rates, and two indicators of whether fixation losses and false negatives were recorded); the eight sector means of RNFL thickness; and the months before the visit. For MD, PSD, visual field index and RNFL thickness the model also read a least-squares slope and a net change over the window, set to zero unless two values spanned at least 180 days. 15 static values described intraocular pressure (the latest reading and its age, means and maxima over the past year and over all prior readings, counts, a slope, and indicators of missing or sparse readings), age and sex. A missing summary was set to zero with a presence flag of zero; the other inputs were standardized on each model’s training rows, with missing values set to zero. The VF encoder pooled features of the 52 total deviation values over eight optic-nerve-head sectors together with MD, PSD, visual field index and a reliability flag, and its training used an auxiliary task that sorted each test’s MD into four bands. Scanning laser ophthalmoscopy images were not used.

Each day’s inputs were projected to 256 numbers per source, combined into one token per day with a continuous-time encoding of its date, and read by a transformer encoder with 2 layers, 4 attention heads, a feed-forward width of 512 and dropout of 0.1; the newest day’s output summarized the window. The trends and then the static values entered through gated layers, and the latest MD and RNFL thickness were added again for the OCT and either-test heads. Three heads (VF, OCT and either test), each with 64 hidden units, gave 20 hazards of 3 months each over 5 years. The risk of progression within a horizon was one minus the product of the survival terms up to it, and the 4-month risk assumed a constant hazard within the second 3-month step.

Each head was trained with a discrete-time survival likelihood on the visits it could judge, censored visits counting as event-free up to their last test, each head with weight 1.0. Two helper tasks predicted the VF indices from the structural inputs and RNFL thickness from the VF input, each with weight 0.3. During training the VF summary, the map summary and the sector means were each hidden at random 20% of the time. Training used AdamW with a learning rate of 0.0003, weight decay of 0.0001, batches of 256 and gradients clipped at a norm of 5, for at most 100 epochs, stopping after 15 epochs without improvement on a held-back eighth of the training patients and keeping the best epoch. Separate models were trained for the entry visit and for each of the next four decision visits, in a four-way rotation over the development folds, so that every development visit was scored by a model that had not seen its patient; each test-set patient was assigned to one rotation model by a fixed hash. Ten runs with different seeds were trained. Their raw risk estimates were averaged on the log-odds scale and then recalibrated once, by logistic regression for each head, horizon and decision visit, pooling over decision visits where a fitting set held fewer than 40 progressions or non-progressions, and made non-decreasing over the horizons by a running maximum. Development visits were recalibrated by fits on the other development folds and test-set visits by fits on all development folds.

### The decision layer and budget selection

The decision layer is specified by a cumulative progression-risk curve for each test, a finite set of candidate intervals, clinical gates defining admissible schedules and penalties for expected testing. It evaluates every admissible schedule and selects the one with the smallest risk-weighted detection delay plus testing penalty. The risk predictor can be changed without changing this decision structure, provided its probabilities refer to the outcomes and horizons used by the decision layer and are appropriately calibrated. In the glaucoma implementation, the two tests were VF and OCT, and the candidate schedules were the 16 interval pairs described below.

For a VF interval v and an OCT interval o, each from {4, 6, 12, 24} months, the decision layer chose the pair that minimized

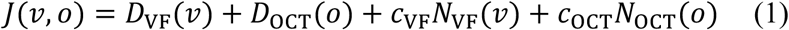

over the admissible pairs. Here *D*_k_(I) is the risk-weighted detection delay of test k (VF or OCT) at interval I, *N*_k_(I) the number of tests of kind k scheduled per year at that interval, repeats included, and *c*_k_ the testing penalty per test, in months. The risk-weighted detection delay is

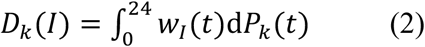

where *P*_k_(t) is the predicted risk that progression becomes detectable on test k within t months, taken as straight lines through (0, 0) and the risks at 3, 4, 6, 12 and 24 months, and *w*_I_(t) is the wait from a change with onset at month t to the next planned test at I, 2I, 3I and so on after the visit. The integral was computed exactly, segment by segment. As intuition, if the risk rose evenly over 2 years, a change would wait half an interval on average, so *D*_k_(I) = *P*_k_(24)I/2.

Equation (2) applies the same idea to the shape of each eye’s predicted risks, so all five horizons shape every interval’s score. The score is additive across modalities, but the VF/OCT interval pair is selected jointly from the pairs permitted by the clinical gates. As a worked example with a hypothetical eye whose risk of VF progression rose evenly to 2% at 24 months (mild stage), *D*_VF_ was 0.04, 0.06, 0.12 and 0.24 months at 4, 6, 12 and 24 months, the testing penalty for a year’s VFs with their repeats was 0.32, 0.21, 0.11 and 0.05 months, and the sums 0.36, 0.27, 0.23 and 0.29 made 12 months the VF interval. At a hypothetical risk of 6% the sums became 0.44, 0.39, 0.47 and 0.77 months, and the interval 6 months. Supplementary Fig. 6 and Supplementary Table 10 show further hypothetical eyes.

The expected tests per year were *N*_VF_(I) = (12/I)(1 + a + *a*_2_) and *N*_OCT_(I) = (12/I)(1 + *a*_OCT_). Here a is the share of routine VFs followed by a confirmation repeat, measured on development visits by the MD stage of the previous VF (0.127, 0.246 and 0.163 for MD above −6 dB, −6 to −12 dB and below −12 dB); *a*_2_ = a(1 − f)(1 − p) is the expected second VF repeat, counted only where twice the repeat delay is shorter than the interval, with f = 0.861 the share of possible VF changes that were not confirmed and p = 0.455 the probability that a first VF repeat confirms; and *a*_OCT_ = 0.036 for an OCT able to judge the eye and zero otherwise.

The ratio of the OCT testing penalty to the VF testing penalty was chosen from 13 values from 1/8 to 8, in equal steps on the log scale. For each ratio, the VF testing penalty was found by 60 halvings on the log scale between 0.001 and 1,000 months as the smallest penalty that scheduled at most 2.00 routine tests per eye per year, averaged over the development scored visits. The budget was 2 routine tests per eye per year, one annual VF and one annual OCT, with repeats on top. The ratio chosen was the one under which GEAR confirmed progression soonest relative to annual testing on development patients in the interval-onset replay (0.7071), giving *c*_VF_ = 0.0943 and *c*_OCT_ = 0.0667 months per test. Exact ties between pairs went to fewer routine tests, then to the shorter VF interval. The budget constrained the development cohort’s mean routine-test count; it was not a per-eye cap or a guarantee of the same aggregate count in a different population. The objective included expected repeat counts, while the development budget excluded repeats. To describe how the budget governed testing (Fig. 3c), both testing penalties were scaled together, with their ratio fixed, over 41 values of the VF testing penalty from 0.01 to 100 months in equal steps on the log scale; at each value we recorded the routine-test rate over all scored visits and the confirmation delay. The perfect-foresight value is the testing rate that would remain at visits without observed progression if GEAR’s total testing rate were kept, every visit followed by progression or operation received its largest admissible testing rate and visits that neither modality could adjudicate kept GEAR’s rate.

The gates applied in this order. At the VF floor (a latest reliable MD of −20 dB or worse) the VF was set at 12 months and the OCT at 12 months or less. At the OCT floor (a latest scan of 57 µm or less, or an OCT unable to judge the eye, with a baseline of 67 µm or less or none) the VF was set at 12 months or less, and an OCT unable to judge the eye at 24 months. The two-year safeguard allowed both tests at 24 months only if *P*_either_(24) − *P*_either_(12) was at most 0.0151, the largest excess at which the development visits allowed to wait 2 years kept a mean excess of at most 0.01, and the OCT was not at its floor. No recommendation was made when the latest VF on or before the visit was not reliable, fewer than two reliable VFs existed, both tests were at their floor, the latest reliable VF or the latest OCT was more than 2 years old or absent, or the ten risk predictor runs, each applying the two-year safeguard to its own recalibrated risks, split five to five.

A possible change on the visit’s VF was a reliable VF on which at least one eligible progression method was abnormal while the confirmation criterion was not yet met; on the OCT it was a scan meeting the COAST threshold that awaited its confirming scan. The delay of the repeat was set at every visit and was 1 month when the visit’s 6-month either-test risk was at least 0.0144 (the development median among visits with a possible VF change), and 2 months otherwise. A repeat followed any test showing a possible change, at the visit or at a later planned test. A second VF repeat followed at twice the delay if the first did not confirm and it fitted before the next planned VF. Confirming a suspected change before acting is standard. Most first abnormal fields in the Ocular Hypertension Treatment Study were not confirmed on retest [17], trials and the perimeter’s progression software require repeated agreement [18,19], and consensus advice is to repeat the field sooner than scheduled when progression is suspected [20]. The second VF repeat is a second opportunity to confirm a real change sooner, not the stricter rule those sources use to avoid false alarms. Each recommendation carried one reason per test, naming the gate that set it or the side of the objective that decided it.

### Timing of the next tests

Every schedule’s next test of each kind fell one interval after the visit, whether or not that test was done at the visit, and the later tests followed at the same interval. A repeat did not move the planned tests.

### Comparators

Annual testing planned the VF and the OCT every 12 months. The guideline-informed comparator, based on the American Academy of Ophthalmology, NICE and European Glaucoma Society guidance [21–23], planned the VF every 12 months in mild and moderate eyes and every 6 months in severe eyes, and every 4 months when intraocular pressure was above 20, 17 or 12 mmHg in mild, moderate or severe eyes, the European target levels by stage standing in for each eye’s own target, which the records do not hold; it planned the OCT every 12 months, or every 6 months in severe eyes. A missing pressure counted as at or below the line. The reconstructed Kalman comparator (with annual OCT) timed the VF by the method of Schell et al. and Helm et al. [5,6], reconstructed on development patients. A Kalman filter held MD, PSD and intraocular pressure, each with its speed and acceleration, in steps of 2 months, with parameters estimated by expectation-maximization on 18,278 development eyes. Helm’s logistic function of MD, its speed and acceleration, PSD, the first PSD and age, fitted by generalized estimating equations to the study’s VF progression on development eyes, turned the forecast into a risk. The next VF came at the first interval on the menu at which the highest risk inside the forecast’s confidence region of size 0.5 reached a threshold of 0.278, set on development visits to schedule 1.00 routine VF per eye per year, as many as annual testing; otherwise it came at 24 months. The cited filtered-forecasting implementations did not schedule OCT, so the reconstructed VF schedule was paired with annual OCT. Supplementary Table 11 lists its settings and the changes from the papers. Every comparator received a confirmation repeat 2 months after a possible change, with the same second VF repeat.

### Dating the onset of progression

The observed-onset replay placed the onset at the first test showing progression. The interval-onset replay spread the onset evenly over the gap from the last test without the change to the first test showing it, at 32 points at (j − 1/2)/32 of the gap (j = 1 to 32) for each test, the OCT’s points in a fixed random order per eye. A visit whose gap ran past the end of its 2-year window counted by the share of its points inside the window, and a gap reaching back before the visit counted in full, an onset before the visit being seen at the first planned test after it (Supplementary Fig. 7). For the worst placement we tried, for each eye, the 32 points and the first test showing the change, and kept the one that gave GEAR its smallest gain over that comparator, the same for both schedules of a comparison. For the uneven onset, some months of the gap were up to 1.5, 2, 3, 4 or 8 times as likely as others.

### Measures

The main measure was the mean months from the onset of progression to its confirmed detection over the visits followed by progression, weighted in the interval-onset replay as above. Months sooner was the comparator’s mean minus that of GEAR, on the same visits. Tests per eye per year were each schedule’s expected count per year, 12/I times one plus the expected repeats for each test, averaged over all scored visits; they do not depend on the replay. We also report the shares of visits followed by progression at which progression was confirmed within 6 months of its onset.

### Hindsight benchmark

The hindsight benchmark took, for each visit, the pair of intervals from the same 16 pairs, under the same gates, timed from the visit and with the visit’s own repeat, that confirmed that eye’s progression soonest in the replay; at visits not followed by progression it took the pair with the fewest tests the gates allow. It holds one plan per visit until the clock ends.

### Statistical analysis

We report 95% CIs from 1,000 bootstrap resamples of patients, drawn with replacement with all of a patient’s eyes and visits together, as percentile intervals; each comparison between schedules was paired on the same resampled visits. The 1,681 test-set patients of the scored visits formed the resampling units. The risk predictor and the frozen settings were not refitted within resamples, and the uncertainty in the measured confirmation probabilities was not carried into the intervals. One 95% level was used for every comparison, with no adjustment for multiplicity; no hypothesis test or interaction test was performed.

### Robustness checks

The worst placement and the uneven onset are described above. For the shuffled-risk check, the predicted risks, the two-year safeguard’s inputs and the repeat’s risk were permuted between visits with a recommendation within the development set and within the test set, 20 times; the floors, the OCT’s ability to judge and the repeat rates stayed with each eye, the same decision layer and testing penalties were applied, and the results were averaged. Even spacing planned both tests at one interval for every eye, chosen so that it scheduled as many tests as GEAR, with a 2-month repeat (12.1 months). The less effective repeat, the shuffled risks and even spacing used the observed-onset replay; the worst placement and the uneven onset are built on the interval-onset replay’s points.

### Learned-policy comparator

To test whether a learned mapping from risk to intervals would schedule better than the explicit objective, we built a learned-policy comparator. A gradient-boosted tree model (Poisson loss, constrained to be non-decreasing in both intervals) predicted the interval-onset confirmation delay of each admissible interval pair in replay. Its inputs were those available to the decision layer (the 15 recalibrated risks, the latest reliable MD, the latest and baseline RNFL thickness, the floor flags, whether the OCT could judge the eye and the months since each test) and the two intervals. It was trained on the development scored visits, with hyperparameters chosen by patient-grouped five-fold cross-validation. Its testing penalties were selected by the procedure used for GEAR, applied to out-of-fold predictions, to meet the same routine-testing budget. In a post hoc stability check, each policy was refitted five times with one fold of development patients withheld, and the pairs chosen at the 9,154 test-set visits with a recommendation were compared with those of the frozen policies. For alternative budgets, the penalties of GEAR were re-selected on the development set by the same procedure.

### Later-period evaluation

For the later-period evaluation, the risk predictor (ten runs) and its recalibration were refitted on development visits up to 31 December 2018, with progression counted only if confirmed by 31 December 2020, and the no-recommendation rule, the repeat-risk threshold and the two-year safeguard were refitted on the same development data. The testing penalties, the gates, the confirmation probabilities of the repeat and the image encoders were not refitted, and whether a test can judge an eye used each eye’s whole follow-up. The reconstructed Kalman comparator (with annual OCT) was refitted the same way, its filter, its progression function and its threshold on development data up to 2018. The test set’s visits from 1 January 2021 were scored. This is a check in later years at the same hospital, not an external validation.

### External VF-only evaluation

The University of Washington’s public dataset [12] holds 28,913 VFs; we kept the 7,398 eyes with at least two, one VF per day, with entry at the second VF and no reliability filter, because the dataset holds no reliability indices. MD was the mean of the 52 total deviation values and PSD their standard deviation. The VF-only adaptation, GEAR-VF, used a Cox model trained on our development patients on the latest MD and PSD, the MD slope, the number of prior VFs, the days since the last VF, the MD at entry and age, recalibrated, giving risks at 6, 12 and 24 months with straight lines between them. It kept the VF terms of the delay-testing objective, the VF floor and the confirmation rule. Worsening was confirmed by at least 2 of 4 of our VF methods (AGIS, the MD trend, pointwise linear regression and PoPLR), in eyes whose starting MD was above −20 dB. The guideline-informed comparator followed MD stage alone (12, 12 and 6 months), the reconstructed Kalman comparator forecast MD and PSD only, with a confidence region of 0.95 and a threshold set to annual testing’s number of VFs on our development visits, and every comparator had the same repeat.

### Software

The analyses used Python 3.13.2, PyTorch 2.6.0, NumPy 2.4.4, pandas 2.2.3, SciPy 1.16.3, scikit-learn 1.8.0, statsmodels 0.14.6, lifelines 0.30.3 and Matplotlib 3.10.1, and the pointwise regression followed the visualFields package [24].

## Data Availability

All data produced in the present study are available upon reasonable request to the authors

